# Cross-sectional associations between prenatal maternal per- and poly-fluoroalkyl substances and bioactive lipids in three Environmental influences on Child Health Outcomes (ECHO) cohorts

**DOI:** 10.1101/2023.11.03.23297930

**Authors:** Himal Suthar, Tomás Manea, Dominic Pak, Megan Woodbury, Stephanie M. Eick, Amber Cathey, Deborah J. Watkins, Rita S. Strakovsky, Brad A. Ryva, Subramaniam Pennathur, Lixia Zeng, David Weller, June-Soo Park, Sabrina Smith, Erin DeMicco, Amy Padula, Rebecca C. Fry, Bhramar Mukherjee, Andrea Aguiar, Sarah Dee Geiger, Shukhan Ng, Gredia Huerta-Montanez, Carmen Vélez-Vega, Zaira Rosario, Jose F. Cordero, Emily Zimmerman, Tracey J. Woodruff, Rachel Morello-Frosch, Susan L. Schantz, John D. Meeker, Akram Alshawabkeh, Max T. Aung, Program Collaborators for Environmental Influences on Child Health Outcomes

## Abstract

**Background:** Per- and poly-fluoroalkyl substances (PFAS) exposure can occur through ingestion of contaminated food and water, and inhalation of indoor air contaminated with these chemicals from consumer and industrial products. Prenatal PFAS exposures may confer risk for pregnancy-related outcomes such as hypertensive and metabolic disorders, preterm birth, and impaired fetal development through intermediate metabolic and inflammation pathways.

**Objective:** Estimate associations between maternal pregnancy PFAS exposure (individually and as a mixture) and bioactive lipids.

**Methods:** Our study included pregnant women in the Environmental influences on Child Health Outcomes Program: Chemicals in our Bodies cohort (CiOB, n=73), Illinois Kids Developmental Study (IKIDS, n=287), and the ECHO-PROTECT cohort (n=54). We measured twelve PFAS in serum and 50 plasma bioactive lipids (parent fatty acids and eicosanoids derived from cytochrome p450, lipoxygenase, and cyclooxygenase) during pregnancy (median 17 gestational weeks). Pairwise associations across cohorts were estimated using linear mixed models and meta-analysis. Associations between the PFAS mixture and individual bioactive lipids were estimated using quantile g-computation.

**Results:** PFDeA, PFOA, and PFUdA were associated (*p*<0.05) with changes in bioactive lipid levels in all three enzymatic pathways (cyclooxygenase [n=6 signatures]; cytochrome p450 [n=5 signatures]; lipoxygenase [n=7 signatures]) in at least one combined cohort analysis. The strongest signature indicated that a doubling in PFOA corresponded with a 24.3% increase (95% CI [7.3%, 43.9%]) in PGD2 (cyclooxygenase pathway) in the combined cohort. In the mixtures analysis, we observed nine positive signals across all pathways associated with the PFAS mixture. The strongest signature indicated that a quartile increase in the PFAS mixture was associated with a 34% increase in PGD2 (95% CI [8%, 66%]), with PFOS contributing most to the increase.

**Conclusions:** Bioactive lipids were revealed as biomarkers of PFAS exposure and could provide mechanistic insights into PFAS’ influence on pregnancy outcomes, informing more precise risk estimation and prevention strategies.

## 1. Introduction

Widespread environmental contamination of per- and poly-fluoroalkyl substances (PFAS) poses a major public health concern. Human exposure to PFAS can occur through ingestion of contaminated food and water, as well inhalation of indoor air from contaminated by consumer products (e.g., clothing, furniture, cookware) (Sunderland et al., 2019). Biomonitoring in the National Health and Nutrition Examination Study (NHANES) has reported over 98% detection of multiple PFAS compounds in serum among individuals in the United States. (Calafat et al., 2007). Importantly, PFAS metabolism and excretion is slow due to their strong carbon-fluorine bonds, with half-lives in the human body spanning from several months to years (Chiu et al., 2022). Increasing experimental and epidemiological evidence indicates adverse health effects attributable to PFAS exposure, including liver toxicity, kidney dysfunction, hormone disruption, and reproductive and developmental toxicity (Fenton et al., 2021). Additionally, biomonitoring of PFAS in NHANES and the Environmental influences on Child Health Outcomes Program indicates moderate correlation between individual compounds, highlighting the need to consider PFAS as a mixture to evaluate cumulative health effects and ameliorate residual confounding (Calafat et al., 2007, Padula et al. 2023).

Pregnancy is a sensitive period of the life course, during which prenatal PFAS exposures have been identified as potential risk factors for adverse birth outcomes and pregnancy complications alike. For example, a systematic review of prenatal PFAS exposures identified select compounds associated with increased odds of preterm birth and miscarriage (Gao et al., 2021). There is also evidence indicating disparities in associations between PFAS exposure in women and chronic health outcomes. For example, a previous study identified that Black women (median age 49) had higher risk of developing hypertension compared to White women (Ding et al. 2023). Our team also identified increased depressive symptoms in association with higher PFAS exposure among immigrant pregnant women compared to U.S. born pregnant women (median age 34) in the Chemicals in Our Bodies cohort (Aung et al. 2023). Widespread evidence of environmental and human exposure, coupled with associated health effects, warrants detailed investigation into intermediate mechanisms of PFAS toxicity to inform risk assessment and identify potential intervention targets.

Bioactive lipids, including poly-unsaturated fatty acids such as arachidonic acid are metabolized by conserved families of enzymes (e.g. cytochrome p450s, lipoxygenases, and cyclooxygenases) to yield secondary eicosanoid metabolites with important downstream physiological functions (Yuan et al., 2018). For example, eicosanoids partly regulate inflammation and influence cardiovascular and renal function, and perturbations in circulating eicosanoids may be important biomarkers of adverse pregnancy outcomes (Eek et al., 2012; Ma et al., 2017; Patel et al., 2004). In a previous metabolomic study in Atlanta, Georgia, lipid metabolism pathways were associated with gestational age at birth, including bioactive lipid compounds such as linoleic acid (Taibl et al., 2023). We have also shown in a previous study in the LIFECODES cohort that several eicosanoid metabolites were associated with increased risk of spontaneous preterm birth (Aung et al., 2019). Another study in the LIFECODES cohort identified eicosanoid metabolites from the cytochrome p450 and lipoxygenase pathway associated with greater risk of being born small for gestational age (Welch et al., 2020).

A review of PFAS exposures and non-targeted metabolomics in epidemiologic studies identified substantial evidence that PFAS are associated with changes to several biological pathways, including metabolism of bioactive lipids, amino acids, and xenobiotic detoxification (Guo et al., 2022). These findings are supported by experimental studies indicating that PFAS can interfere with cytochrome p450 signaling, which is critical for cellular metabolism (Hvizdak et al., 2023; Zanger and Schwab, 2013). PFAS can also interfere with homeostasis of intracellular calcium gradients, which in turn can impact calcium dependent enzymatic activity and alter systemic bioactive lipid profiles (Cao and Ng, 2021). Given the increasing evidence that PFAS are linked to whole pathways of lipid metabolism, there is a need for deeper investigation of targeted lipid metabolites to determine precise molecular signatures of PFAS exposure and implications for pregnancy outcomes and fetal development.

The objective of this study was to quantify maternal PFAS exposures and bioactive lipids during pregnancy and estimate the individual and cumulative associations of PFAS on these biomarkers. There is a need for modern epidemiology studies to integrate greater racial, socioeconomic, and geographic diversity to better inform risk estimation across historically marginalized communities and for varied contexts of PFAS exposure levels. Thus, in the current study, we utilized a diverse study sample across three birth cohorts in the Environmental influences on Child Health Outcomes Program. Our study hypothesis was that higher concentrations of PFAS are associated with increased bioactive lipid signatures across enzymatic groups that govern eicosanoid synthesis.

## 2. Methods

### 2.1 Study Populations

The Environmental influences on Child Health Outcomes (ECHO) Program comprises 69 ongoing and new pregnancy and pediatric cohorts. The ECHO Program’s goal is to advance research that increases understanding of how environmental factors spanning from preconception through childhood influences child health and development (Knapp et al. 2023). This study integrates data across three ECHO cohorts (**Table 1**): Chemicals in Our Bodies (CiOB), Illinois Kids Development Study (IKIDS), and the ECHO-PROTECT cohort (Eick et al., 2021; Ferguson et al., 2019). The CiOB cohort participants are based in San Francisco, CA, and were recruited during the second trimester of pregnancy at three University of California San Francisco hospitals. Women included in the CiOB cohort had to be at least 18 years of age, speak English or Spanish as a primary language, and be having a singleton pregnancy. The IKIDS cohort participants were recruited between 10 and 14 weeks’ gestation from two obstetric clinics in Champaign-Urbana, Illinois. Women included in the IKIDS cohort had to be between 18 and 40 years of age, have English fluency, have a singleton pregnancy, be ≤ 15 weeks gestation at enrollment, not have a child already in the IKIDS cohort, reside within a 30 minute drive from the University of Illinois Urbana-Campaign (UIUC) campus, and plan to remain in the area until at least the child’s first birthday. The ECHO-PROTECT cohort participants were recruited before 20 weeks’ gestation from two hospitals and five clinics in the Northern Karst aquifer region in Puerto Rico. Enrollment of the cohort began in 2011 and is ongoing. Inclusion criteria for the women in the ECHO-PROTECT cohort included being between 18 and 40 years of age, residing in the Northern Karst aquifer region, not have used oral contraceptives during the three months pre-pregnancy, not have undergone in-vitro fertilization, and have no major pre-existing conditions. Detailed information on study recruitment and data collection on sociodemographic and health information for each cohort has been previously described (Eick et al., 2021; Ferguson et al., 2019). Participants in each cohort provided written informed consent to be included in this study and local institutional review boards for each cohort reviewed and approved study protocols. Sample sizes for each cohort are described in greater detail below, and the flow diagram for selection of the final analytical dataset used in statistical analyses is illustrated in **Supplemental Figure 1**.

**Table 1.** Demographic profile of CiOB, IKIDS, and ECHO-PROTECT cohorts.

| Characteristic |  | Cohort |  |  |  | *p* value<sup>b</sup> |
| --- | --- | --- | --- | --- | --- | --- |
|  |  | Overall | CiOB, N = 73<sup>a</sup> | IKIDS, N = 287<sup>a</sup> | ECHO-PROTECT, N = 54<sup>a</sup> |  |
| Mother's Race |  |  |  |  |  | <0.001 |
|  | Non-Hispanic White | 253 (62%) | 34 (47%) | 219 (78%) | 0 |  |
|  | Black | 19 (54.6%) | - | 14 (5.0%) | 0 |  |
|  | Asian | 48 (12%) | 14 (19%) | 34 (12%) | 0 |  |
|  | Hispanic | 78 (19%) | 17 (23%) | 7 (2.5%) | 54 (100%) |  |
|  | Other | 11 (2.7%) | - | 8 (2.8%) | 0 |  |
|  | Missing | 5 | 0 | 5 | 0 |  |
| Maternal age (years) |  | 31.8 (28.8, 34.5) | 33.1 (29.9, 36.0) | 31.8 (29.1, 34.3) | 28.0 (24.0, 33.0) | <0.001 |
|  | Missing | 6 | 1 | 0 | 5 |  |
| Maternal education |  |  |  |  |  | <0.001 |
|  | < HS | 11 (2.7%) | - | - | - |  |
|  | HS/GED/some college | 77 (19%) | 16 (22%) | 39 (14%) | 22 (45%) |  |
|  | Bachelors | 142 (35%) | 15 (21%) | 111 (39%) | 16 (33%) |  |
|  | Graduate | 179 (44%) | 37 (51%) | 135 (47%) | 7 (14%) |  |
|  | Missing | 5 | 0 | 0 | 5 |  |
| Pre-pregnancy BMI |  | 25 (22, 29) | 24 (22, 27) | 25 (22, 30) | 25 (21, 29) | 0.2 |
|  | Missing | 20 | 11 | 1 | 8 |  |
| Parity |  |  |  |  |  | <0.001 |
|  | 0 | 191 (47%) | 35 (51%) | 156 (54%) | 0 (0%) |  |
|  | 1 or more | 214 (53%) | 34 (49%) | 131 (46%) | 49 (100%) |  |
|  | Missing | 9 | 4 | 0 | 5 |  |
| Maternal household |  |  |  |  |  | <0.001 |
| | <\$50,000 | 110 (28%) | 18 (26%) | 52 (18%) | 40 (93%) | |
| | \$50,000-\$99,999 | 145 (36%) | 6 (9%) | 137 (48%) | - | |
| | >\$100,000 | 143 (36%) | 45 (65%) | 97 (34%) | - | |
|  | Missing | 16 | 4 | 1 | 11 |  |
| Gestational age at visit |  | 17.4 (16.6, 18.9) | 23.1 (19.7, 25.7) | 17.0 (16.4, 17.7) | 25.7 (24.4, 27.9) | <0.001 |
|  | Missing | 10 | 0 | 0 | 10 |  |

<sup>a</sup>n (%), cells with counts ≤ 5 have been masked; Median (IQR)<sup>b</sup>Kruskal-Wallis rank sum test; Pearson's Chi-squared test

### 2.2. PFAS Exposure Assessment

In the CiOB cohort (n = 73, median gestational age of sample collection = 23 weeks) and IKIDS (n = 287, median gestational age of sample collection = 17 weeks) cohorts, twelve PFAS compounds were quantified in maternal serum during pregnancy: Perfluorononanoic acid (PFNA), Perfluoroheptanoic acid (PFHpA), Perfluorodecanoic acid (PFDeA), Perfluorododecanoic acid (PFDoA), Perfluorooctanoic acid (PFOA), Perfluorooctanesulfonamide (PFOSA), 2-(N-Methyl-perfluorooctane sulfonamido) acetic acid (Me-PFOSA-AcOH), 2-(N-Ethyl-perfluorooctane sulfonamido) acetic acid (Et-PFOSA-AcOH), Perfluoroundecanoic acid (PFUdA), Perfluorohexane sulfonate (PFHxS), Perfluorooctanesulfonic acid (PFOS), and Perfluorobutanesulfonic acid (PFBS). After sample collection, serum was frozen at −80°C. Samples were processed at the Environmental Chemical Laboratory at the California Department of Toxic Substances Control using a previously described analytical protocol (Morello-Frosch et al., 2016). PFAS quantification was achieved using internal standards in each serum sample and an automated on-line solid phase extraction method coupled to liquid chromatography and tandem mass spectrometry. The limits of detection for individual PFAS compounds were equal to three times the standard deviation of the blank negative control sample (Morello-Frosch et al., 2016). In each batch of serum PFAS analyses, analytes were quantified using a constructed calibration, and regression coefficients (R^2^) of 0.98 to 0.99 were generally obtained. We utilized in-house quality control materials that were prepared by spiking a known amount of PFAS compounds in blank bovine serum at low and high levels. We utilized standard reference materials (SRM 1958) from the National Institute of Standards and Technology (Gaithersburg, MD), and quality control samples spiked with known PFAS concentrations from the United States Centers for Disease Control and Prevention as reference materials. Blank samples of bovine serum (Hyclone/GE Healthcare LifeSciences) were also processed with each batch of samples, and no PFAS were detected above their respective limited of detection in these blank samples.

Maternal serum samples from the ECHO-PROTECT cohort (n = 54, median gestational age of sample collection =26 weeks) were used to quantify nine PFAS compounds (PFNA, PFHpA, PFDeA, PFOA, PFOSA, Me-PFOSA-AcOH, PFUdA, PFHxS, PFOS). After sample collection, serum was frozen at −80°C and processed at NSF International (Ann Arbor, MI, USA). PFAS quantification was also performed by liquid chromatography and tandem mass spectrometry. The method simulates the Centers for Disease Control and Prevention’s polyfluoroalkyl chemicals laboratory procedure method No: 6304.1. Standards of known purity and identity were used during preparation of the calibration, quality control, and internal standards. The validated analyte calibration curve correlation coefficient (R^2^) ranges were 0.996 or greater. The method accuracy (% nominal concentration) and precision (% relative standard deviation [RSD]) were determined through six replicate analyses of analytes spiked at three different concentrations in pooled human serum across validation runs on three separate days (n = 18), which reflects both the intra-day and inter-day variability of the assay. The accuracy (% nominal concentration) range across all analytes was 95.1–103% with precision (%RSD) range for the serum quality control samples across all analytes being 2.3–16%.

For samples with values below the limit of detection, we used machine read values (if reported) or concentrations imputed using the limit of detection divided by the square root of 2 (Hornung and Reed, 1990). Of the PFAS compounds measured, we selected to analyze those with a detection rate of ≥ 70%, which included PFNA, PFHpA, PFDeA, PFOA, Me-PFOSA-AcOH, Et-PFOSA-AcOH, PFUdA, PFHxS, and PFOS in the CiOB cohort; PFNA, PFDeA, PFOA, Me_PFOSA-AcOH, PFUdA, PFHxS, and PFOS in the IKIDS cohort; and PFNA and PFOS in the ECHO-PROTECT cohort (**Table 2A**).

**Table 2A.** PFAS detection rates and distributions across cohorts.

| PFAS (ng/mL) |  | CiOB, N = 73 |  | IKIDS, N = 287 |  |  | ECHO-PROTECT, N = 54 |  |  | *p* value<sup>b</sup> |
| --- | --- | --- | --- | --- | --- | --- | --- | --- | --- | --- |
| Full name<br>(abbreviation) | MDL<sup>a</sup> | %<br>Detected | Median<br>(IQR) | MDL<sup>a</sup> | %<br>Detected | Median<br>(IQR) | MDL<sup>a</sup> | %<br>Detected | Median<br>(IQR) |  |
| Perfluorononanoic acid (PFNA) | **0.04; 0.03** | **99%** | **0.53 (0.34, 0.72)** | **0.03** | **100%** | **0.40 (0.26, 0.59)** | **0.1** | **81%** | **0.20 (0.16, 0.29)** | <0.001 |
| Perfluoroheptanoic acid (PFHpA) | **0.04; 0.02** | **82%** | **0.03 (0.02, 0.05)** | 0.02 | 58% | 0.03 (0.015, 0.03) | 0.1 | 0% | 0.10 (0.10, 0.10) | - |
| Perfluorodecanoic acid (PFDeA) | **0.06; 0.04** | **96%** | **0.17 (0.11, 0.29)** | **0.04** | **98%** | **0.11 (0.07, 0.19)** | 0.1 | 31% | 0.10 (0.10, 0.16) | <0.001 |
| Perfluorododecanoic acid (PFDoA) | 0.14; 0.04 | 66% | 0.06 (0.02, 0.16) | 0.08 | 55% | 0.10 (0.01, 0.11) | - | - | - | - |
| Perfluorooctanoic acid (PFOA) | **0.04** | **99%** | **1.13 (0.66, 1.92)** | **0.04** | **100%** | **1.03 (0.62, 1.73)** | 0.5 | 41% | 0.50 (0.50, 0.96) | 0.4 |
| Perfluorooctanesulfonamide (PFOSA) | 0.02; 0.01 | 66% | 0.01 (0.004, 0.02) | 0.01 | 55% | 0.01 (0.009, 0.01) | 0.1 | 0% | - | - |
| 2-(N-Methyl-perfluorooctane sulfonamido) acetic acid (Me-PFOSA-AcOH) | **0.01** | **93%** | **0.06 (0.02, 0.09)** | **0.008** | **95%** | **0.07 (0.03, 0.15)** | 0.1 | 0% | - | 0.07 |
| 2-(N-Ethyl-perfluorooctane sulfonamido) acetic acid (Et-PFOSA-AcOH) | **0.009; 0.008** | **71%** | **0.01 (0.006, 0.01)** | 0.008 | 40% | 0.01 (0.01, 0.01) | - | - | - | - |
| Perfluoroundecanoic acid (PFUdA) | **0.04; 0.02** | **96%** | **0.18 (0.11, 0.29)** | **0.02** | **91%** | **0.06 (0.03, 0.12)** | 0.1 | 24% | 0.10 (0.10, 0.10) | <0.001 |
| Perfluorohexane sulfonate (PFHxS) | **0.01** | **99%** | **0.55 (0.33, 0.98)** | **0.06** | **99%** | **0.76 (0.40, 1.42)** | 0.1 | 50% | 0.12 (0.10, 0.17) | 0.016 |
| Perfluorooctanesulfonic acid (PFOS) | **0.04** | **99%** | **2.96 (1.88, 5.52)** | **0.04** | **100%** | **3.27 (2.01, 5.28)** | **0.1** | **100%** | **1.78 (1.40, 2.28)** | 0.8 |
| Perfluorobutanesulfonic acid (PFBS) | 0.02 | 48% | 0.03 (0.02, 0.03) | 0.02 | 40% | 0.03 (0.01, 0.03) | - | - | - | - |

Bolded measurements denote PFAS with detection rates $\geq 70\%$ and used in analyses.<sup>a</sup>Method Detection Limits (ng/mL).<sup>b</sup>Kruskal-Wallis Rank Sum Test; performed only across cohorts which had $\geq 70\%$ detection rates for specific PFAS.

### 2.3. Bioactive Lipids Assay

Maternal plasma samples were used to quantify a targeted panel of 50 bioactive lipids (**Table 2B)** in the CiOB (n = 73, median gestational age of sample collection = 24 weeks), IKIDS (n = 287, median gestational age of sample collection = 17 weeks), and PROTECT (n = 54, median gestational age of sample collection = 26 weeks) cohorts. For ECHO-PROTECT and CiOB cohorts, plasma was collected using ethylenediaminetetraacetic acid plasma tubes and temporarily stored at +4°C for less than four hours. Blood was subsequently centrifuged for 20 minutes and stored at −80°C. For participants in the IKIDS cohort, women provided a fasted (10 to 12 hours) blood sample collected in green-top sodium heparin tubes. IKIDS participant blood samples were kept at room temperature for two hours prior to processing and centrifuged at room temperature for 20 minutes. The resulting plasma was aliquoted immediately into cryovials for storage at −80°C. The targeted bioactive lipids panel consisted of five parent fatty acid compounds and 45 eicosanoid metabolites derived from three enzymatic groups (lipoxygenases, cytochrome p450s, and cyclooxygenases), and full names and abbreviations are documented in **Table 2B**. Quantification of bioactive lipid concentrations was achieved using a 6490 Triple Quadrupole mass spectrometer (Agilent, New Castle, DE, USA). In this setting, the mass spectrometer was set to a targeted multiple reaction monitoring mode and individual biomarkers were identified based on metabolite-specific fragmentation and retention time. Limits of detection were not calculated across individual instrument analysis cycles and all measured bioactive lipids had machine read values. Further information on instrumental parameters and quality control/assurance have been previously documented (Afshinnia et al., 2020). Briefly, we conducted sequential dilution of each internal standard in duplicate to establish linearity and estimate the coefficient of variation (CV) of the measurements at various concentrations. We also ran a pool of study samples at the beginning of each batch and then after each 12 samples during mass spectrometry to assess drift in measurements over time as well as the batch-to-batch variability.

**Table 2B.** Bioactive lipids sampled from maternal plasma.

| Pathway | Full name | Abbreviation |
| --- | --- | --- |
| Cyclooxygenase | 13,14-dihydro-15-keto Prostaglandin F2α | 13,14DHK-PGF2a* |
| Cyclooxygenase | 13,14-Dihydro-15-keto Prostaglandin J2 | 13,14-D-PGJ2 |
| Cyclooxygenase | 15-deoxy-Δ12,14-Prostaglandin J2 | 15DO12,14-PGJ2 |
| Cyclooxygenase | Bicyclo Prostaglandin E2 | BCPGE2 |
| Cyclooxygenase | Bicyclo Prostaglandin E1 | BCPGE1 |
| Cyclooxygenase | Prostaglandin A2 | PGA2 |
| Cyclooxygenase | Prostaglandin B2 | PGB2 |
| Cyclooxygenase | Prostaglandin D2 | PGD2 |
| Cyclooxygenase | Prostaglandin D3 | PGD3 |
| Cyclooxygenase | Prostaglandin E1 | PGE1 |
| Cyclooxygenase | Prostaglandin E2 | PGE2 |
| Cyclooxygenase | Prostaglandin E3 | PGE3 |
| Cyclooxygenase | Prostaglandin J2 | PGJ2 |
| Cyclooxygenase | Thromboxane B2 | TXB2 |
| Cyclooxygenase | 9-Oxo-octadeca-dienoic acid | 9-oxoODE |
| Cytochrome p450 | 20-carboxy Arachidonic Acid | CAA |
| Cytochrome p450 | 9,10-Dihydroxy-octadecenoic acid | 9,10-DiHOME |
| Cytochrome p450 | 12,13-Dihydroxy-octadecenoic acid | 12,13-DiHOME |
| Cytochrome p450 | 13,14-dihydro-15-keto Prostaglandin D2 | 13,14DHK-PGD2 |
| Cytochrome p450 | 5,6-Dihydroxy-eicosatrienoic acid | 5,6-DHET |
| Cytochrome p450 | 8,9-Dihydroxy-eicosatrienoic acid | 8,9-DHET |
| Cytochrome p450 | 11,12-Dihydroxy-eicosatrienoic acid | 11,12-DHET |
| Cytochrome p450 | 9,10-Epoxy-octadecenoic acid | 9(10)-EpoME |
| Cytochrome p450 | 12,13-Epoxy-octadecenoic acid | 12(13)-EpoME |
| Cytochrome p450 | 5,6-Epoxy-eicosatrienoic acid | 5(6)-EET |
| Cytochrome p450 | 8,9-Epoxy-eicosatrienoic acid | 8(9)-EET |
| Cytochrome p450 | 11,12-Epoxy-eicosatrienoic acid | 11(12)-EET |
| Cytochrome p450 | 14,15-Epoxy-eicosatrienoic acid | 14(15)-EET |
| Cytochrome p450 | 11-Hydroxy-eicosatetraenoic acid | 11-HETE |
| Cytochrome p450 | 16-Hydroxy-eicosatetraenoic acid | 16-HETE |
| Cytochrome p450 | 17-Hydroxy-eicosatetraenoic acid | 17-HETE |
| Cytochrome p450 | 18-Hydroxy-eicosatetraenoic acid | 18-HETE |
| Cytochrome p450 | 20-Hydroxy-eicosatetraenoic acid | 20-HETE |
| Cytochrome p450 | 9S-Hydroxy-octadecadienoic acid | 9S-HODE |
| Lipoxygenase | 13S-Hydroxy-octadecadienoic acid | 13S-HODE |
| Lipoxygenase | 5-Hydroxy-eicosatetraenoic acid | 5-HETE |
| Lipoxygenase | 8-Hydroxy-eicosatetraenoic acid | 8-HETE |
| Lipoxygenase | 12-Hydroxy-eicosatetraenoic acid | 12-HETE |
| Lipoxygenase | 15-Hydroxy-eicosatetraenoic acid | 15-HETE |
| Lipoxygenase | 5-Oxo-eicosatetraenoic acid | 5-oxoETE |
| Lipoxygenase | 12-Oxo-eicosatetraenoic acid | 12-oxoETE |
| Lipoxygenase | 15-Oxo-eicosatetraenoic acid | 15-oxoETE |
| Lipoxygenase | 13-Oxo-octadeca-dienoic acid | 13-oxoODE |
| Lipoxygenase | Resolvin D1 | RVD1 |
| Lipoxygenase | Resolvin D2 | RVD2 |
| Parent Compound | α-Linolenic Acid | αLA |
| Parent Compound | Arachidonic Acid | AA |
| Parent Compound | Docosahexaenoic Acid | DHA |
| Parent Compound | Eicosapentaenoic Acid | EPA |
| Parent Compound | Linoleic Acid | LA |

### 2.4. Statistical Analyses

We utilized a conceptual model (**Figure 1**) to inform our statistical analyses. We tabulated distributions of key covariates from our conceptual model for each of the cohorts in our study. Spearman correlations were estimated between all high-detect (i.e., ≥ 70% of observations exceeded minimum level of detection in cohort) PFAS compounds and bioactive lipids within each of the cohorts. Combined-cohort correlations were not conducted due to differences in high-detect PFAS across cohorts. Based on which PFAS were highly detected in each cohort (**Table 2A**), associations were estimated in combined cohort samples with two (CiOB and IKIDS) or three cohorts (CiOB, IKIDS, and ECHO-PROTECT) using linear mixed effect regressions (random intercept for cohort), and meta-analysis. Significance was evaluated at a level of α ≤ 0.05. Associations between PFAS mixtures and bioactive lipids were estimated using quantile g-computation in the combined cohort consisting of IKIDS and CiOB participants (Keil et al., 2020). All analyses are described in greater detail below and were performed with R (version 4.3.0).

**Figure 1.**
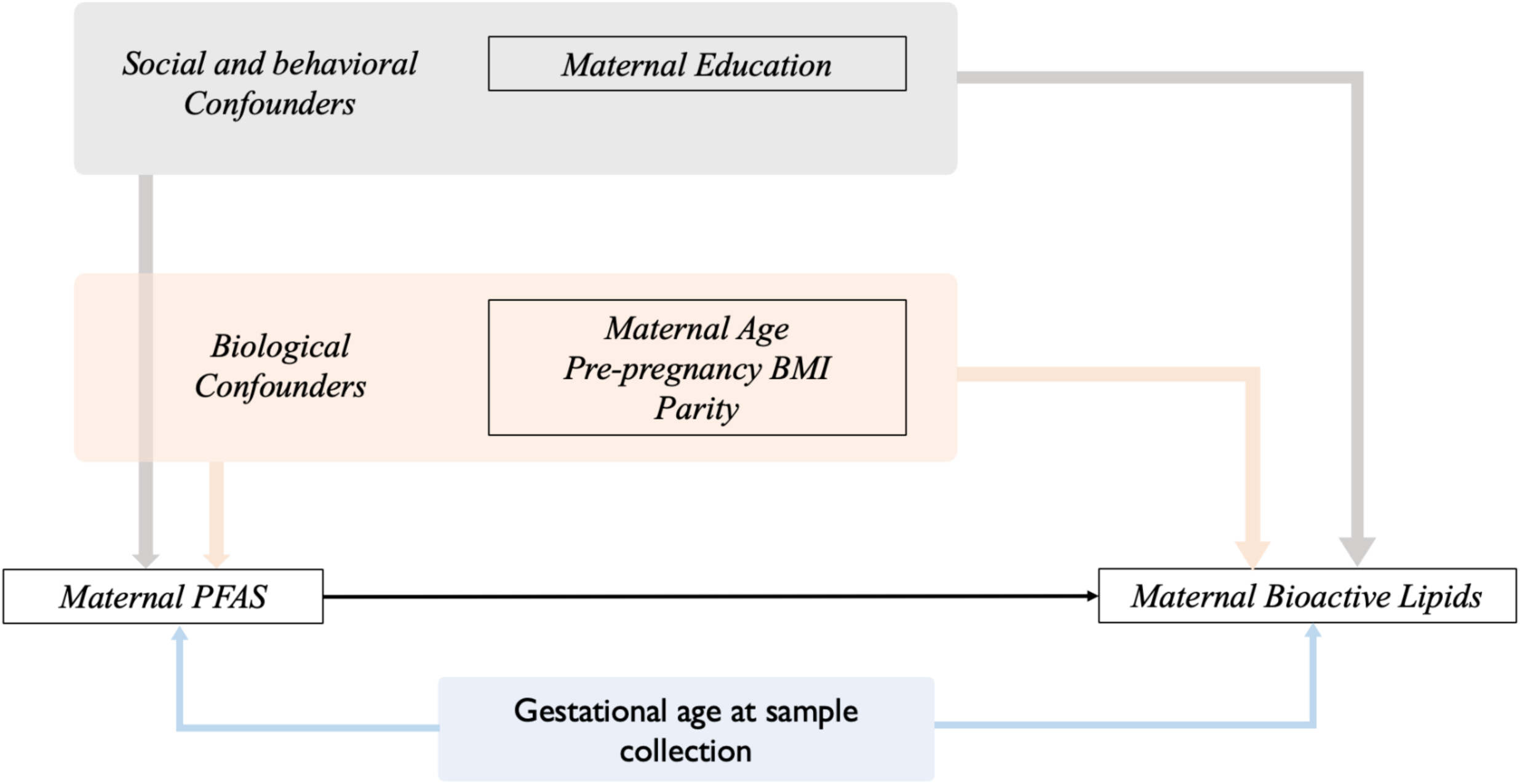
Directed acyclic graph of the relationships between maternal per- and poly-fluoroalkyl substances and bioactive lipids

#### 2.4.1. Combined cohort analysis

Linear mixed effects models were utilized to test combined-cohort pairwise associations between all sampled bioactive lipids and common high-detect PFAS, using natural log-transformed lipid levels as outcomes and natural log-transformed PFAS as exposures. These linear mixed effects models included a random intercept for the cohort identity to partly ameliorate bias from different laboratory analysis of PFAS and for heterogeneity in demographic characteristics between cohorts. All effect estimates were back-transformed to enhance interpretability and represent a percent change in a given bioactive lipid corresponding to a doubling (or 100% increase) of individual PFAS. Unadjusted and adjusted random intercept models were generated for all three cohorts using PFAS common to all three cohorts: PFNA and PFOS (**Table 2A**). Unadjusted and adjusted random intercept models were also generated combining the CiOB and IKIDS cohorts, using the high-detect PFAS common to those two cohorts as exposures: ME-PFOSA-AcOH, PFDEA, PFHxS, PFNA, PFOA, PFOS, and PFUDA (**Table 2A**). Adjusted models included variables selected *a priori* based on our previous investigation of PFAS and biomarkers of lipid peroxidation and oxidative stress (Taibl et al., 2022). The covariates included in the adjusted models were maternal age in years, education (categorized into less than high school, high school/GED/some college, bachelor’s degree, and graduate degree), pre-pregnancy BMI, parity (categorized into 0 and 1 or more), and gestational age in weeks at sampling collection for bioactive lipids. Missing observations for covariates were omitted from adjusted models. Q-values were calculated for all pairwise models to control for false discovery rates (Storey et al., 2015)

#### 2.4.2. Meta-analysis

Pairwise effect estimates for PFAS and bioactive lipids from each individual cohort were estimated using linear regression, adjusted for the same covariates modeled in the combined cohort analysis with the exception of ECHO-PROTECT, which did not include parity as a covariate due to 0% variance in the variable within the ECHO-PROTECT cohort. We combined these effect estimates by meta-analysis using the METAL method (Willer et al., 2010), yielding overall effect estimates and *p* values for pairwise associations between individual bioactive lipids and PFAS. This method, used previously in genome-wide association studies, utilizes an inverse variance calculation to weigh all beta coefficients by their standard errors. In doing so, this approach allows researchers to obtain effect estimates across models with different underlying populations and covariates, without requiring access to the raw data. Meta-analysis was implemented for all three cohort-specific models, and then for the IKIDS and CiOB cohort-specific models. Interpretation of cohort-specific effect estimates were treated as a sensitivity analysis to evaluate consistencies in combined cohort and meta-analysis and reported in supplementary materials.

#### 2.4.3. PFAS mixtures analysis

To address the study goal of estimating the cumulative effect of multiple PFAS on bioactive lipids, we used quantile g-computation (Keil et al., 2020), a generalization of the weighted quantile sum (WQS) regression method (Carrico et al., 2015; Czarnota et al., 2015), which does not assume directional homogeneity of the exposures’ effects on the outcome. This method outputs an unbiased estimate of the effect on a particular bioactive lipid associated with simultaneously increasing all PFAS exposures by one quartile. Quantile g-computation was implemented in the combined CiOB and IKIDS sample to measure the mixtures effect of log-transformed high-detect PFAS common to both the CiOB and IKIDS cohorts (Et-PFOSA-AcOH, PFDEA, PFHxS, PFNA, PFOA, PFOS, and PFUDA; **Table 2A**) on all log-transformed bioactive lipids, while controlling for the same covariates included in the adjusted combined CiOB and IKIDS random intercept model. The cohort itself was included as an additional fixed effect covariate in the models to account for any between-cohort differences (Taibl et al., 2022).

## 3. Results

### 3.1. Descriptive statistics

**Table 1** shows the distribution of maternal race/ethnicity, age, education, pre-pregnancy BMI, parity, household income category, and gestational age at plasma collection for bioactive lipids across all three cohorts. White women represented the largest group in CiOB (47%) and IKIDS (78%), and ECHO-PROTECT had only Hispanic women (100%). Women in CiOB were the oldest with a median age of 33 years, followed by IKIDS (median age = 32 years), and ECHO-PROTECT (median age = 28 years) (*p* < 0.01). Women in IKIDS had the highest educational attainment, with 86% having completed a bachelor’s or graduate degree, followed by CiOB (72%) and ECHO-PROTECT (47%; *p* < 0.01). All mothers in ECHO-PROTECT reported having at least one previous birth, which was much higher compared to CiOB (49%) and IKIDS (46%, *p* < 0.01). The annual household income for women in CiOB was the highest with 65% reporting annual household incomes of ≥ $100,000, followed by women in IKIDS (34%), while 93% of women in ECHO-PROTECT reported a household income < $50,000 (*p* < 0.01). Income was not adjusted for local cost of living or median salary. Pre-pregnancy BMI was similar across all three cohorts (median 24 kg/m^2^ in CIOB and 25 kg/m^2^ in IKIDS and ECHO-PROTECT, *p* = 0.2).

Distributions of PFAS and bioactive lipids by cohort are reported in **Table 2A** and **Supplemental Table 1**, respectively. Among the high-detect PFAS common to all three cohorts, PFNA (median 0.20 ng/mL) and PFOS (median 1.78 ng/mL) concentrations were the lowest in the ECHO-PROTECT cohort, (*p* < 0.01) while the CiOB cohort had the highest median concentration of PFNA (0.53 ng/mL), and the IKIDS cohort had the highest median concentration of PFOS (3.27 ng/mL). Among the high-detect PFAS common to only the CiOB and IKIDS cohorts, the greatest differences were observed for PFUdA (median 0.18 ng/mL in CiOB and median 0.06 ng/mL in IKIDS; *p* < 0.01) and PFHxS (median 0.55 ng/mL in CiOB and median 0.76 ng/mL in IKIDS; *p* = 0.02). Among the bioactive lipids, we observed the greatest difference across cohorts for linoleic acid concentrations (median 668 µMol/L in CiOB, 1,185 µMol/L in IKIDS, and 102 µMol/L in ECHO-PROTECT; *p* < 0.01) and 20-carboxy arachidonic acid (CAA) concentrations (median 185 nMol/L in CiOB, 302 nMol/L in IKIDS, and 74 nMol/L in ECHO-PROTECT; *p* < 0.01).

### 3.2. Within-Cohort Correlations

Within-cohort Spearman correlations between bioactive lipids and PFAS are shown in **Figure 2A** for the CiOB cohort, **Figure 2B** for the IKIDS cohort, and **Figure 2C** for the ECHO-PROTECT cohort. In CiOB, correlation coefficients between bioactive lipids and PFAS ranged between −0.31 and 0.38, with the strongest negative correlation between DHA and PFOS, and the strongest positive correlation between 13S-HODE and PFUDA. For bioactive lipids in the CiOB cohort, correlation coefficients ranged from −0.47 (16-HETE and 13-oxoODE) and 1 (12(13)-EpoME and 13S-HODE), while correlation coefficients for PFAS were between −0.08 (PFDeA and Et-PFOSA-AcOH) and 0.81 (PFNA and PFOS). In IKIDS, correlation coefficients between bioactive lipids and PFAS ranged between −0.14 and 0.22, with the strongest negative correlation between 15-HETE and PFHxS, and the strongest positive correlation between arachidonic acid and PFNA. Correlation coefficients for bioactive lipids in the IKIDS cohort were between −0.75 (13,14-D-PGJ2 and 18-HETE) and 0.89 (9S-HODE and 13S-HODE), and between 0.05 (PFOA and Me-PFOSA-AcOH) and 0.75 (PFNA and PFOA) for PFAS. In ECHO-PROTECT, correlation coefficients between bioactive lipids and PFAS ranged between −0.37 and 0.32, with the strongest negative correlation between 9,10-DiHOME and PFNA, and the strongest positive correlation between 13,14-D-PGJ2 and PFNA. Bioactive lipid correlation coefficients within the ECHO-PROTECT cohort were between 0.59 (PGA2 and 11-HETE) and 0.8 (9(10)-EpoME and 12(13)-EpoME), and the correlation coefficients between PFNA and PFOS in the ECHO-PROTECT cohort was 0.57.

**Figure 2A.**
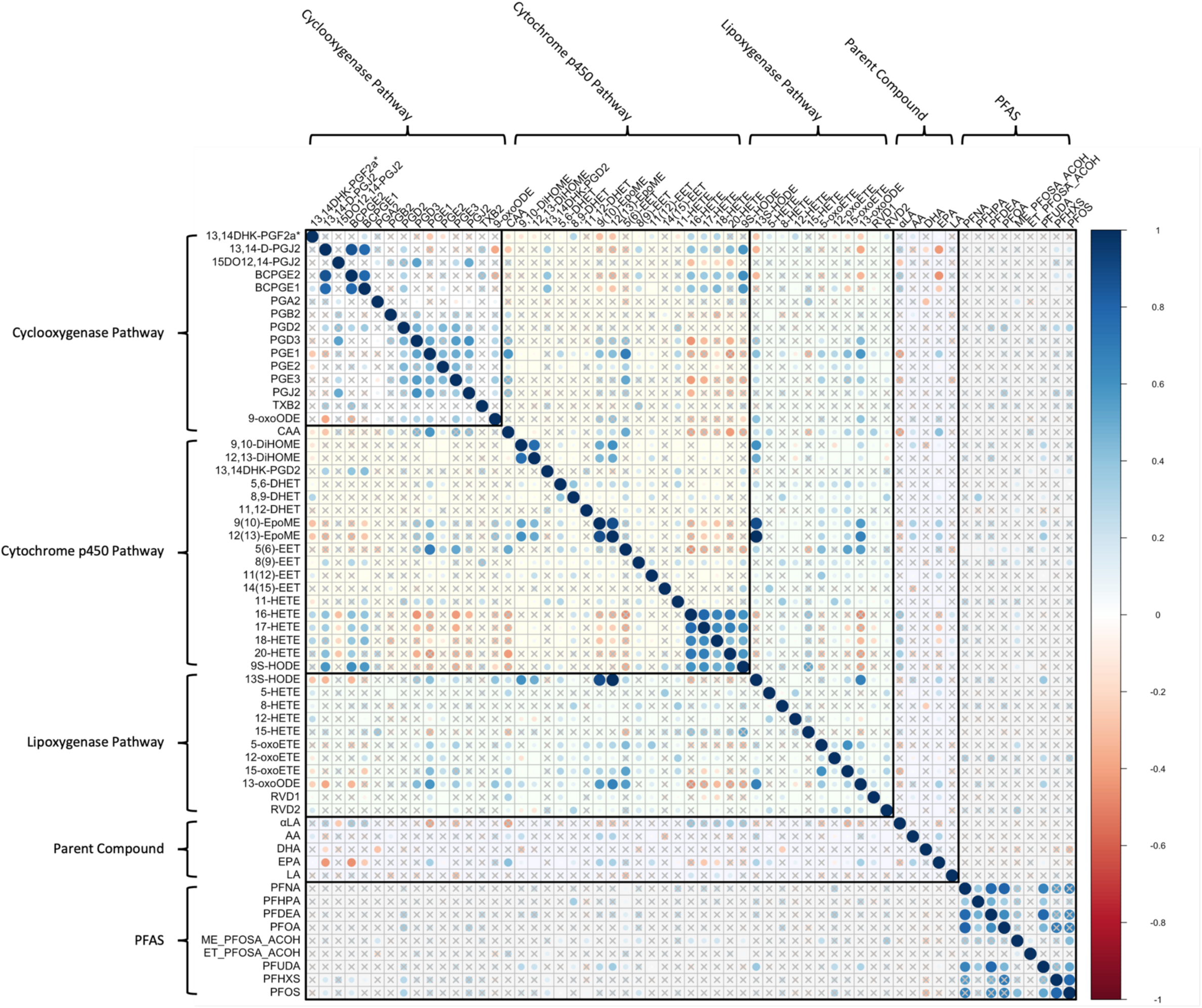
Correlation Matrix of Bioactive Lipids and High-Detect PFAS in the CiOB Cohort

**Figure 2B.**
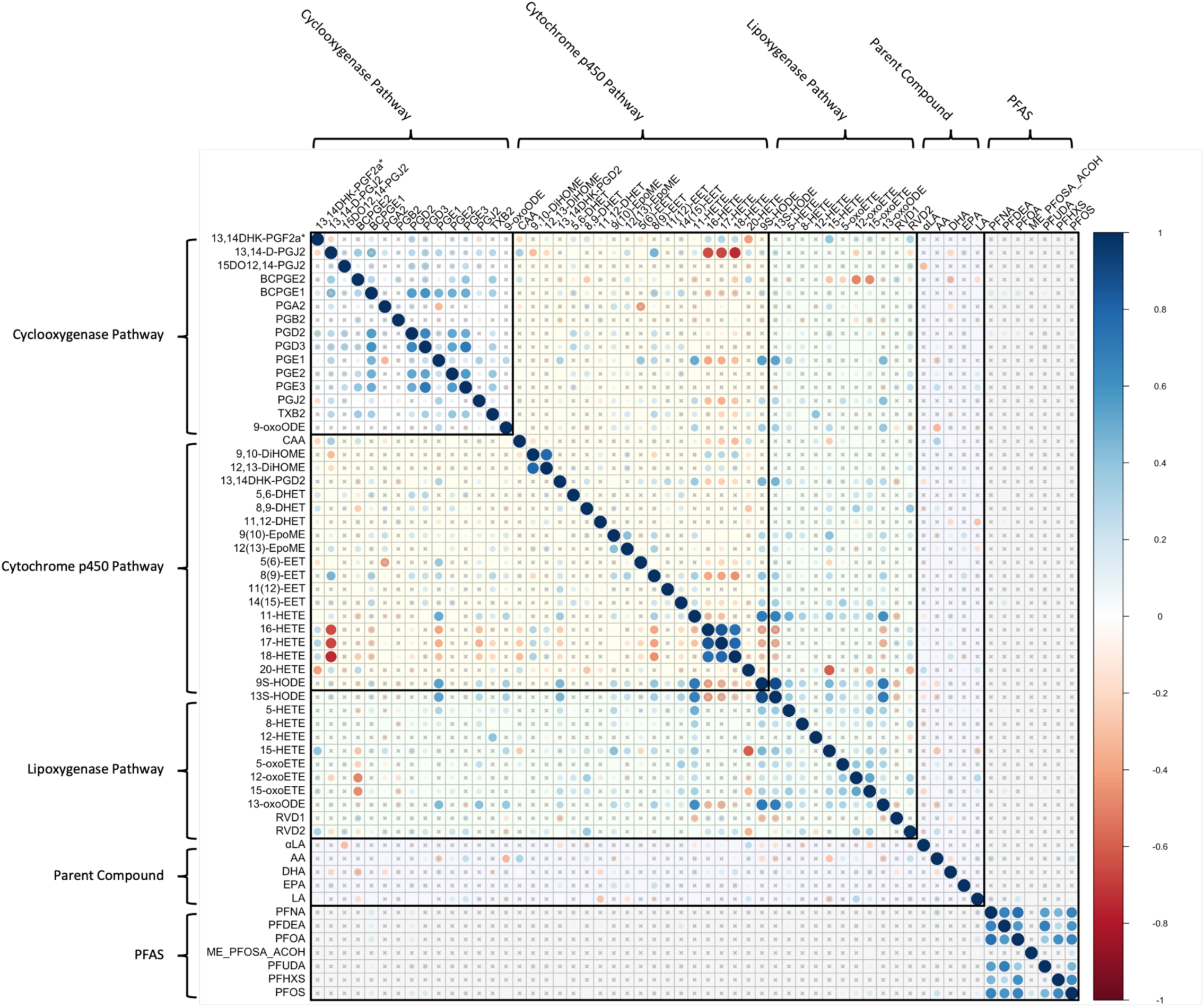
Correlation Matrix of Bioactive Lipids and High-Detect PFAS in the IKIDS Cohort

**Figure 2C.**
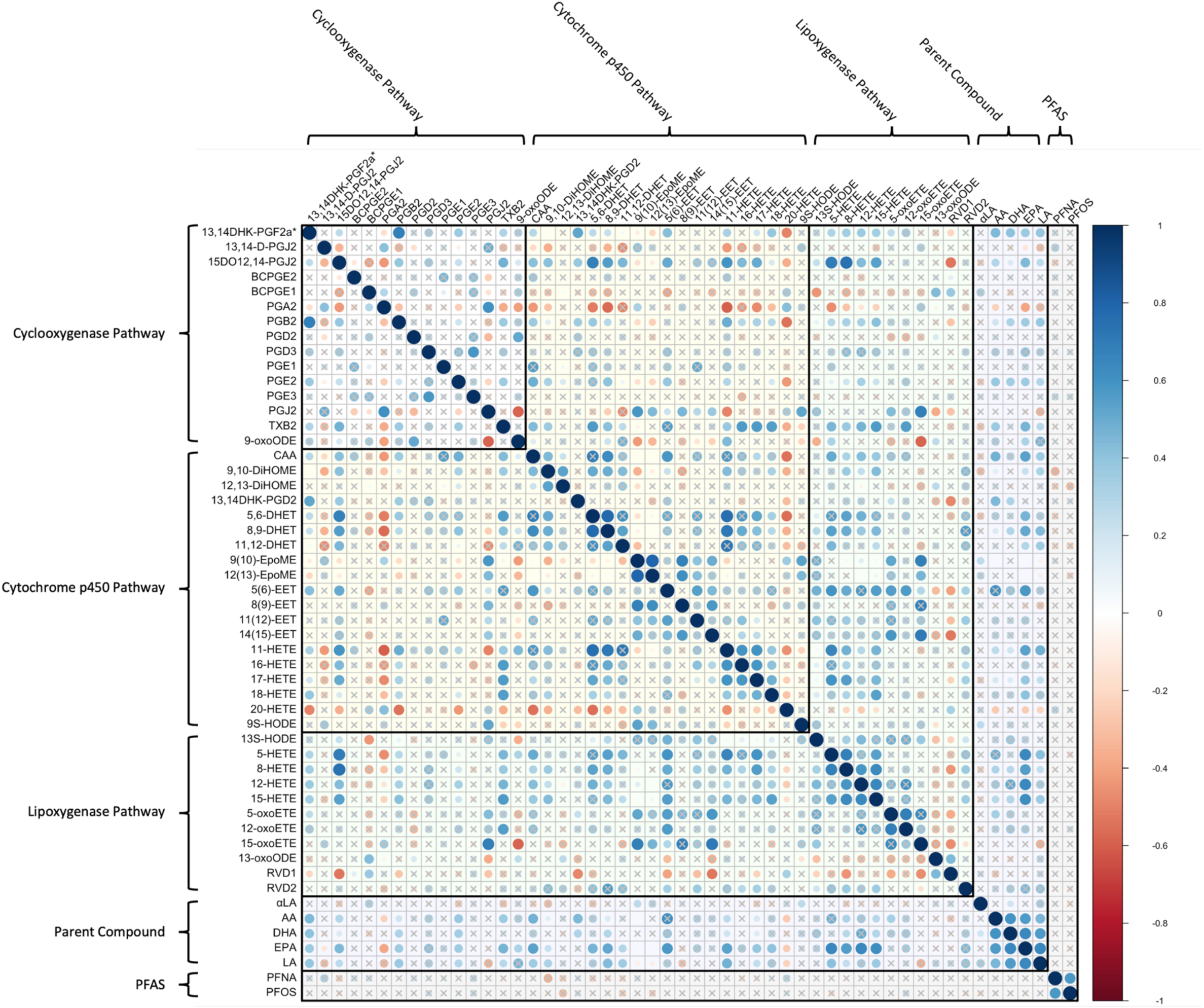
Correlation Matrix of Bioactive Lipids and High-Detect PFAS in the ECHO-PROTECT Cohort

### 3.3. Pair-wise associations between individual PFAS and bioactive lipids

All coefficients representing the pairwise associations between bioactive lipids and PFAS from the adjusted random intercept models and meta-analyses are presented in a heatmap in **Figure 3** as the effect per doubling of PFAS concentration. Additional output of these models, including exact *p* values and standard errors can be found in **Supplemental Tables 2-5**. Additionally, effect estimates and 95% confidence intervals from significant associations found in at least one of the four models are reported in **Table 3**. With the exception of the associations between 9-oxoODE (cytochrome p450 pathway) and PFNA, 15-HETE (lipoxygenase pathway) and PFHxS, and αLA and PFUdA (parent compounds), all significant associations observed in at least one of the four combined models were positive.

**Figure 3.**
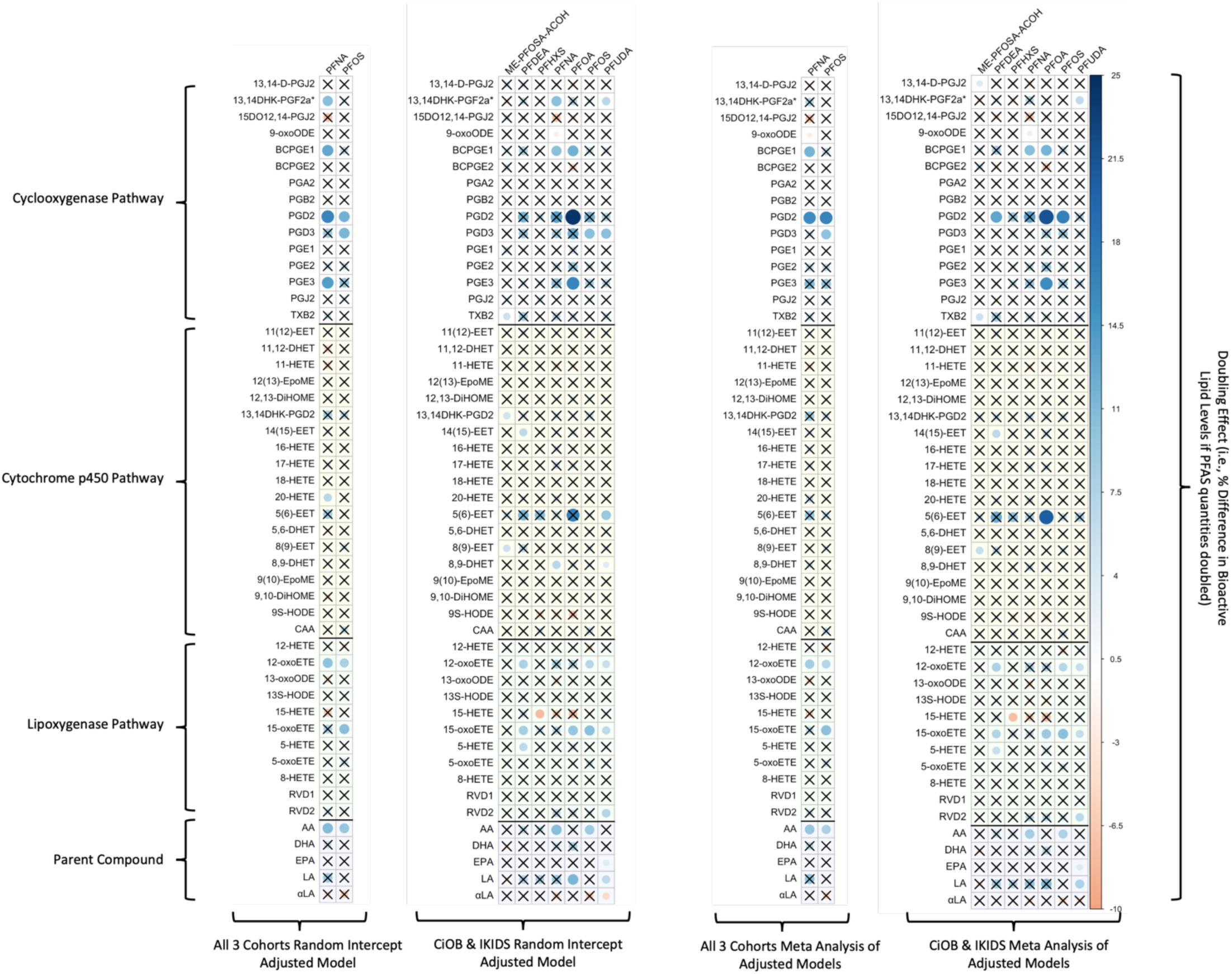
Heatmap of β Estimates Corresponding to Percentage Change in Bioactive Lipids as a Result of Doubling Log-Transformed PFAS for Adjusted Joint Analyses and Meta-Analyses run on Adjusted Within-Cohort Models. The magnitude of effect estimates in each cell in the heatmap corresponds to the intensity of the color band in the legend. Non-significant values (p > .05) are marked with black “X”. Sample sizes range from 343 to 383 (see Supplemental Tables 3, 4, 5, and 12 for exact pair-wise sample sizes).

**Table 3.**
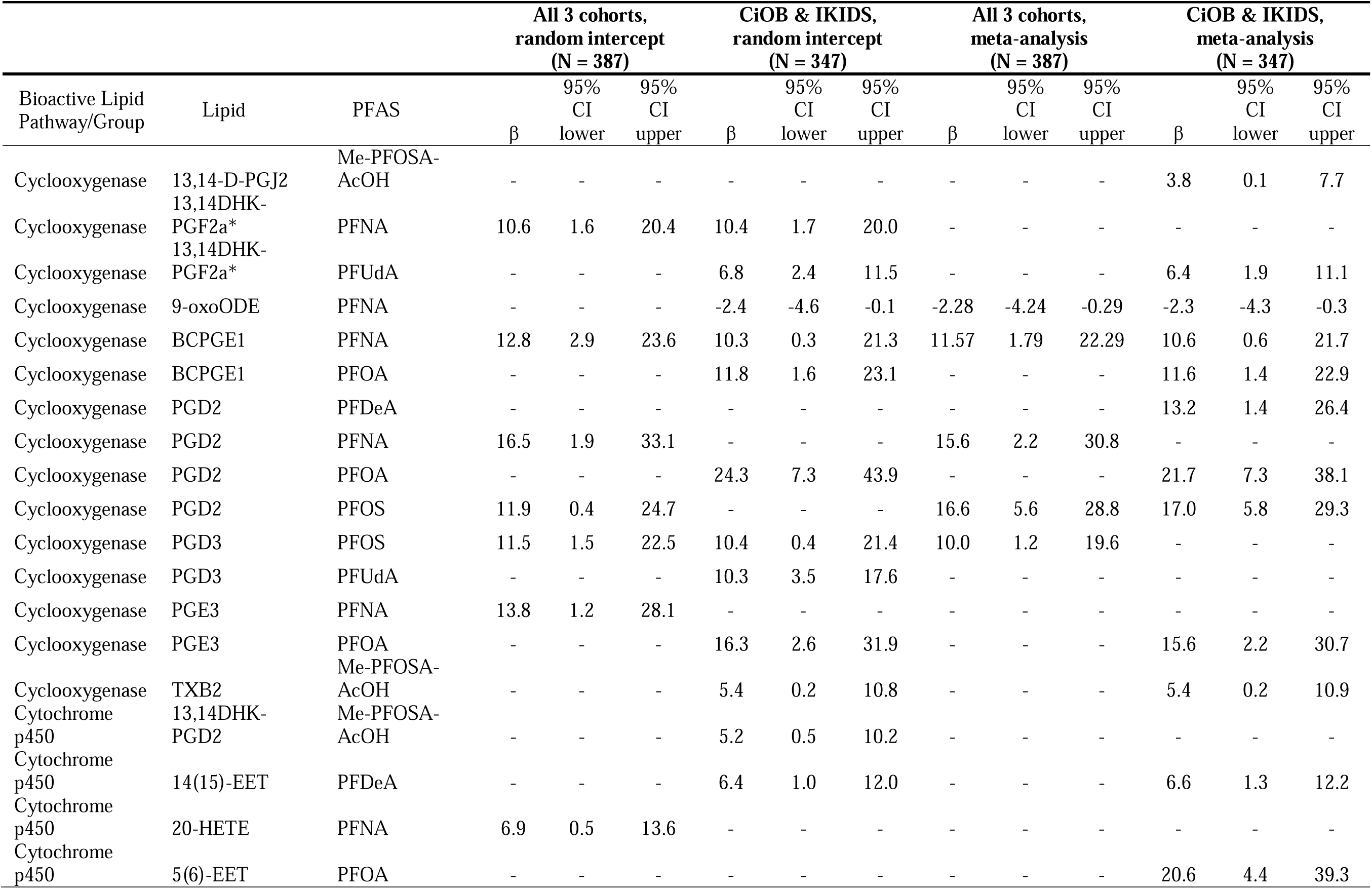
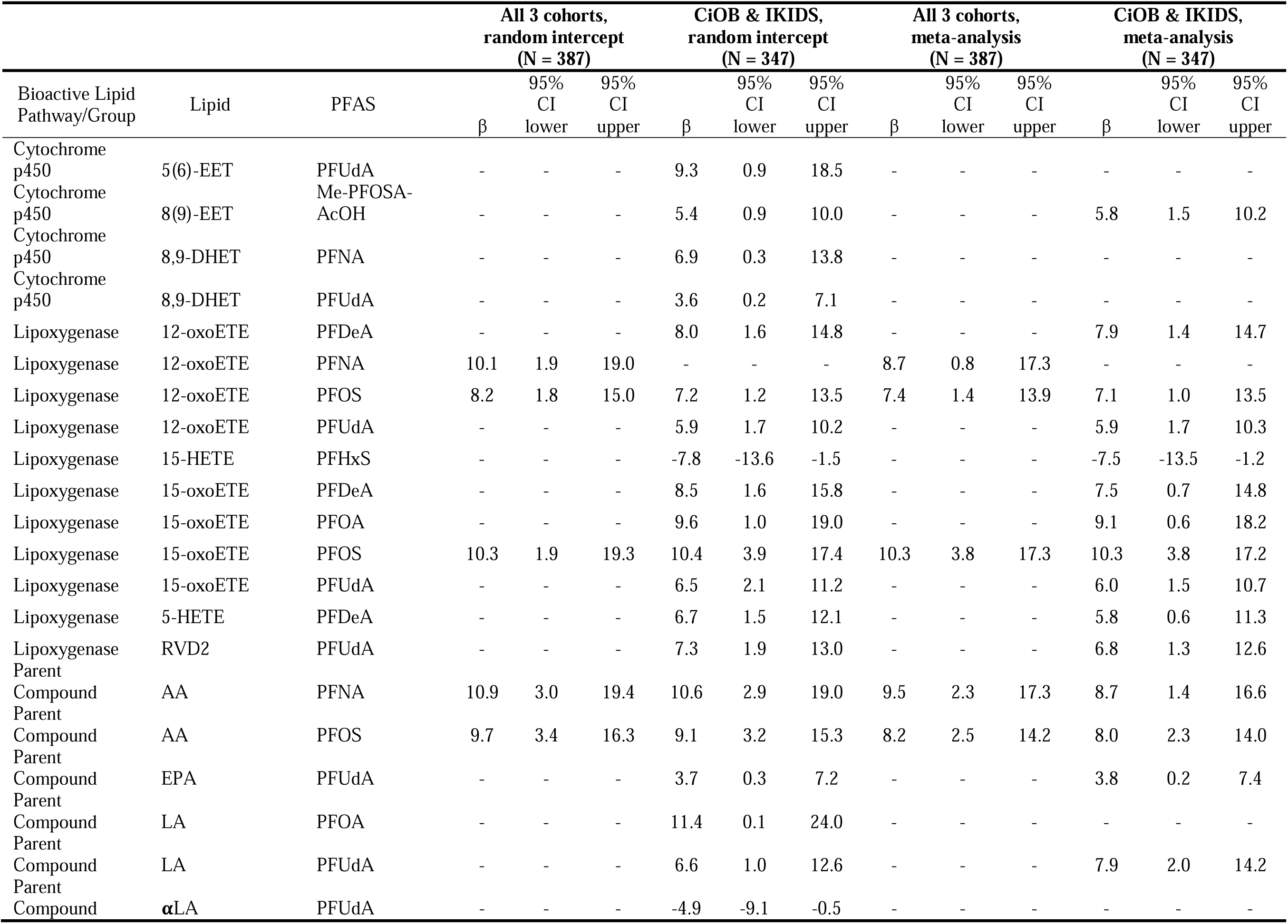
β estimates and 95% confidence intervals corresponding to percentage change in bioactive lipids associated (*p* ≤ 0.05) with a doubling in PFAS; for joint cohort and meta-analyses.

In the cyclooxygenase pathway, 15 significant associations between bioactive lipids and PFAS were observed in at least one of the four combined cohort models. Of these, the association between BCPGE1 and PFNA was significant in all four models (doubling effect in PFNA ranging from 10.3% to 12.8% increase in BCPGE1 across models). Additionally, the associations between 9-oxoODE and PFNA (doubling effect in PFNA ranging from 2.3% to 2.4% decrease in 9-oxoODE), PGD2 and PFOS (doubling effect in PFOS ranging from 11.9% to 17.0% increase in PGD2), and PGD3 and PFOS (doubling effect in PFOS ranging from 10.0% to 11.5% increase in PGD3) were significant in three of the four models. In the cytochrome p450 pathway, no significant associations were found which were common to all four models; however, there were significant positive associations between 14(15)-EET and PFDeA (doubling effect ranging from 6.4% to 6.6%) and between 8(9)-EET and Me-PFOSA-AcOH (doubling effect ranging from 5.4% to 5.8%) observed in the CiOB and IKIDS random intercept model and in the meta-analysis integrating effect estimates from CiOB and IKIDS. Within the lipoxygenase pathway, significant positive associations were observed between 12-oxoETE and PFOS (doubling effect ranging from 7.1% to 8.2%) and 15-oxETE and PFOS (doubling effect ranging from 10.3% to 10.4%) across all four combined cohort models. Among the parent compounds, significant positive associations were observed for arachidonic acid and PFNA (doubling effect ranging from 8.7% to 10.9%) and arachidonic acid and PFOS (doubling effect ranging from 8.0% to 9.7%) across all four combined cohort models.

We explored multiple testing comparison adjustments in the linear mixed effects models (**Supplemental Tables 2 and 3**). While none were below a threshold of 0.1 in the two cohort or three cohort models, there were eight PFAS and bioactive lipid pairs (PGD2 and PFOA; PGD3 and PFUdA; 13,14DHK-PGF2a* and PFUdA; 12-oxoETE and PFUdA; 15-oxoETE and PFUdA; 15-oxoETE and PFOS; arachidonic acid and PFNA; arachidonic acid and PFOS) below the 0.2 threshold in the two cohort models. Our sensitivity analysis of within-cohort pairwise associations can be found in **Supplemental Tables 6-11**. While individual cohort analyses are underpowered to detect associations, directions of pair-wise associations between individual cohorts and combined cohorts were largely consistent.

### 3.4. PFAS Mixture Associations

Quantile g-computation utilizing the CiOB and IKIDS cohorts indicated that simultaneously increasing all log-transformed PFAS (PFNA, PFDeA, PFoA, Me-PFOSA, PFUdA, PFHxS, and PFOS) in the mixture by one quartile corresponded to increases in BCPGE1, PGD2, PGD3, and PGE3 in the cyclooxygenase pathway; 12,13-DiHOME and 5(6)-EET in the cytochrome p450 pathway; 12-oxoETE, 15-oxoETE, and 5-ETE in the lipoxygenase pathway, and arachidonic acid and linoleic acid parent compounds. The largest increase in the cyclooxygenase pathway was observed in PGD2 (34% increase, 95% CI [8%, 66%]), with decomposition of the quantile g-computation effect estimate for PGD2 indicating that PFOS exhibited the greatest positive weight to the overall mixture effect relative to the other PFAS compounds in the overall sample (**Supplemental Table 12**). In the cytochrome p450 pathway, the largest increase associated with a simultaneous one quartile increase in all PFAS in the mixture was in 5(6)-EET (31% increase, 95% CI [5%, 64%]), with PFHxS contributing the most to this increase. The largest increase in the lipoxygenase pathway was observed in 12-oxoETE (16% increase, 95% CI [4%, 30%]), with PFUdA having the largest contribution to this increase. In the parent compounds, linoleic acid had the largest increase (18% increase, 95% CI [1%, 38%]), driven predominantly by PFUdA. No significant decreases in bioactive lipids were observed. These effects are visualized in **Figure 4** and reported in detail in **Supplemental Table 12**.

**Figure 4.**
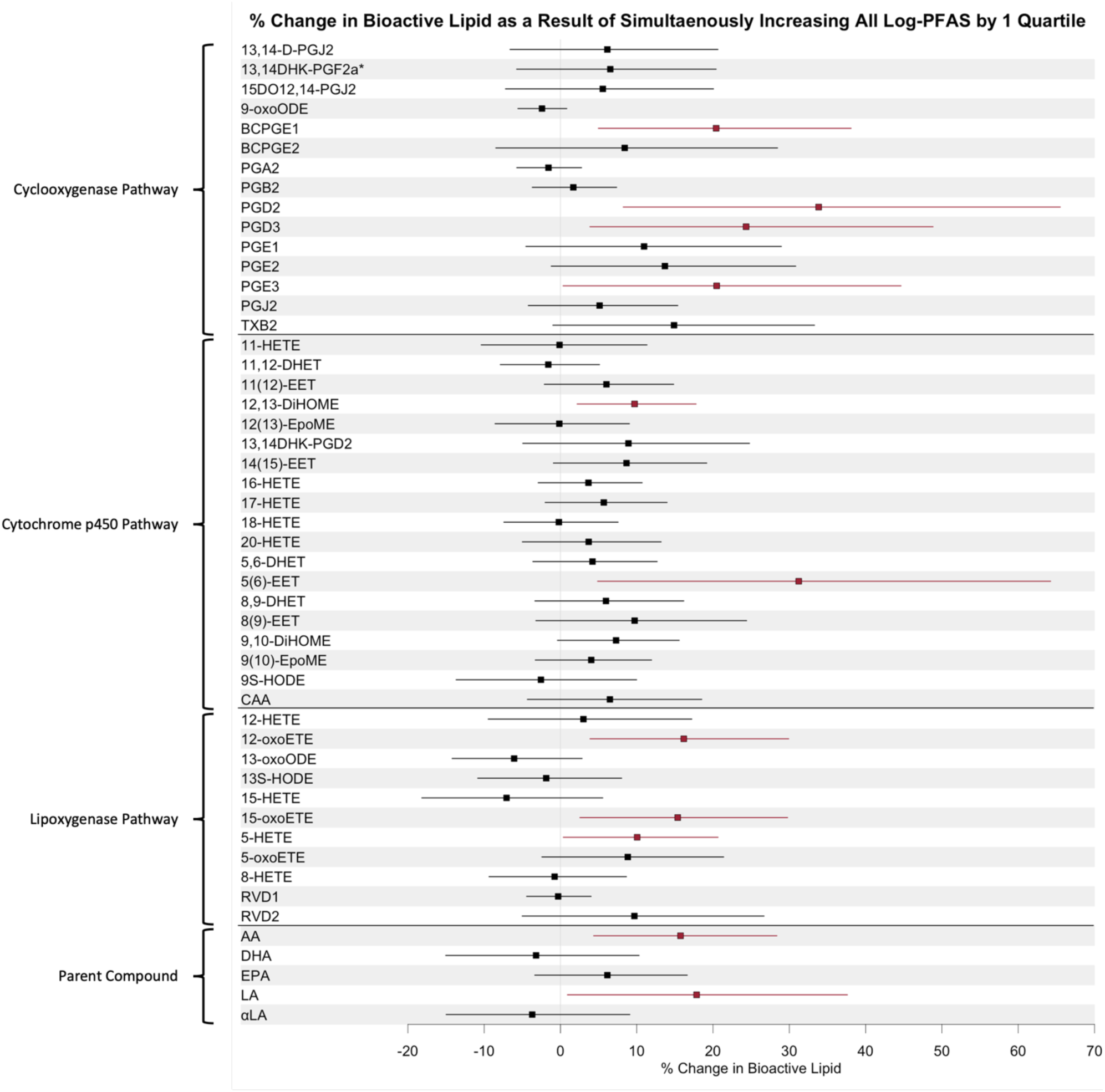
Forest Plot of Quantile g-computation Effect Estimates in the Combined Cohort Analysis with IKIDS and CIOB Cohorts (N=343), showing of β Estimates Corresponding to Percentage Change in Bioactive Lipids as a Result of Simultaneous 1-Quartile Increase in All Log-Transformed PFAS. Model adjusted for Maternal Age, Maternal Education, Pre-Pregnancy BMI, Parity, Gestational Age at Visit, and Cohort.

## 4. Discussion

### 4.1. Summary of findings across statistical approaches

In the present study, we advanced mechanistic insight into PFAS exposures during pregnancy by estimating associations between PFAS and bioactive lipids in single pollutant and mixture models. We utilized multiple statistical approaches to estimate associations, including cohort stratified analyses, combined cohort analyses, and meta-analyses. Combined cohort analyses and meta-analyses identified mostly positive associations of PFAS with parent fatty acid compounds and their secondary eicosanoid metabolites derived from the lipoxygenase, cytochrome p450, and cyclooxygenase pathways. Individual cohorts have limited power to detect significant associations, which underscores the utility of exploring combined cohort analyses and meta-analyses. Additionally, mixtures analysis using quantile g-computation revealed that the entire PFAS mixture exhibited predominantly positive associations with bioactive lipids across all three enzymatic pathways (BCPGE1, PGD2, PGD3, PGE3, 12(13)-DiHOME, 5(6)-EET, 12-oxoETE, 15-oxoETE, 5-HETE, arachidonic acid, and linoleic acid), and, with the exception of 12(13)-DiHOME, at least one individual PFAS was associated with these bioactive lipids in the single pollutant models for either combined cohort analyses or meta-analyses. Largely consistent results between mixtures analysis and pair-wise associations strengthen confidence in targeted bioactive lipids as a potential mechanistic biomarker of PFAS exposure and provide insight into addressing the health effects of PFAS as an entire class. Doing so can inform more precise risk estimation and ameliorate the influence of residual confounding resulting from co-occurring PFAS compounds. Although our estimated associations are susceptible to false-positive associations, our findings still allow for prioritization of pairs of PFAS and bioactive lipids for future hypothesis testing and replication in independent samples.

### 4.2. Biological context of associations in bioactive lipid enzymatic pathways

The physiological implications of our findings vary based on the metabolic pathway under investigation. In our study, we observed the strongest positive effect between the PFAS mixture and cyclooxygenase derived metabolite PGD2, which was consistent with individual PFAS compound analyses for PFDeA, PFOA, and PFOS in the CiOB and IKIDS meta-analyses. Systemic inflammation and oxidative stress are both antecedent physiological states that increase the risk of adverse pregnancy outcomes such as spontaneous preterm birth and preeclampsia (Ferguson and Chin, 2017; Gomez-Lopez et al., 2022). Cyclooxygenases have been implicated in promoting inflammation through the production of prostaglandins (Ricciotti and FitzGerald, 2011). Prostaglandin production is also sensitive to imbalances in reactive oxygen species and oxidative stress (Burdon et al., 2007). Previous mechanistic studies indicate that cyclooxygenases are also important for regulating reproductive health and fetal development, where animal models have found that deficiencies in the genes encoding these enzymes result in altered implantation, increased mortality, and impaired organ development in offspring (Xu et al., 2007). Animal studies indicate that disruptions in cyclooxygenase function and prostaglandin synthesis can also lead to altered neurodevelopment and behavior (Davis-Bruno and Tassinari, 2011; Wong et al., 2019). For example, a previous study in an experimental rat model found that the PGD2 signaling pathway is involved in neuroinflammation, and induction of this pathway results in neurodegenerative pathologies (Corwin et al., 2018). Evidence from previous experimental mechanistic studies in combination with our findings that PFAS are linked to altered eicosanoid concentrations within the cyclooxygenase pathway underscore the need to further investigate this pathway as a potential mediator between PFAS and adverse pregnancy outcomes.

In our study, we observed that the PFAS mixture was associated with two cytochrome p450 derived eicosanoids: 12,13-DiHOME and 5(6)-EET. Cytochrome p450 enzymes have varied regulatory roles, including biosynthesis of endogenous hormones, detoxification of xenobiotics, and cellular metabolism (Zanger and Schwab, 2013). Importantly, *in vitro* experiments indicate that multiple PFAS compounds can directly interfere and inhibit cytochrome p450 activity (Hvizdak et al., 2023). 12,13-DiHOME has been classified as an oxylipin derived from linoleic acid, and is involved with inflammation, endocrine signaling, and adipogenesis (Hildreth et al., 2020; Macêdo et al., 2022). In a previous case-control study that our research team led in the LIFECODES cohort using the same bioactive lipids panel measured (median 26 weeks’ gestation) in this present study, we reported that 12,13-DiHOME was associated with increased risk of spontaneous preterm birth (n_cases_ = 31, n_controls_ = 115) (Aung et al., 2019). Further, placental 5(6)-EET has been detected at higher levels in preeclamptic women compared to controls, linking 5(6)-EET to regulation pathways associated with preeclampsia (Dalle Vedove et al., 2016; Herse et al., 2012). Additionally, a study of 146 adult women reported that single nucleotide polymorphisms of cytochrome p450 genes amplified cancer risk attributable to PFAS exposures (Ghisari et al., 2014). While bioactive lipids were not measured in that study, this is consistent biological inference based on their findings of increased breast cancer risk in association with higher PFOS and PFOA and polymorphisms in cytochrome p450 genes. Therefore, cytochrome p450 derived eicosanoids may link PFAS to adverse maternal health outcomes and serve as early signals to inform precise prevention efforts.

Lipoxygenases are calcium dependent enzymes that catalyze the formation of hydroperoxides from poly-unsaturated fatty acids and have been associated with several adverse health outcomes including asthma, skin disorders, and cancers (Mashima and Okuyama, 2015). In our study, we observed that the PFAS mixture was associated with higher levels of the lipoxygenase derived eicosanoids 12-oxoETE, 15-oxoETE, and 5-HETE. These oxylipins may be important biomarkers for metabolic and cardiovascular disorders. For example, 12-oxoETE has been explored as a biomarker of diabetic macular edema (Rhee et al., 2021) and experimental models indicate that 15-oxoETE may influence atherosclerosis (Ma et al., 2017). Additionally, our previous study in the LIFECODES cohort identified higher levels of 12-oxoETE and 5-HETE in association with spontaneous preterm birth (Aung et al., 2019). Our present study findings showing that increases in 12-oxoETE and 15-oxoETE were associated with increased PFAS exposure are further aligned with previous experimental evidence showing that PFAS can interfere with intracellular calcium gradients which can influence the catalytic activity of lipoxygenases (Cao and Ng, 2021). Therefore, future studies should continue to investigate this pathway as a mechanistic link between PFAS exposure and adverse maternal and child health outcomes.

We observed differences in bioactive lipid compound distributions between individual cohorts, which may be driven by the variation in genetic makeup of the populations caused by heterogeneous ethnic composition of the cohorts (Sergeant et al., 2012), or differences in environmental exposures of the cohorts due to diet (Saadatian-Elahi et al., 2009). Among the parent poly-unsaturated fatty acids, the PFAS mixture was associated with increased concentrations of arachidonic acid and linoleic acid. These findings are aligned with the positive signatures we observed for the secondary eicosanoid metabolites described above that are derived from these parent compounds. Our reported findings can partially be contextualized with previous metabolomics studies to confer biological inference of PFAS-induced effects on lipid metabolism. One metabolomics study investigated a mixture of six common PFAS (PFOS, PFHxS, PFHpS, PFOA, PFNA, and PFDA) in children and adolescents (n = 137) based in Los Angeles, CA, and observed positive associations of the mixture with arachidonic acid and linoleic acid (Goodrich et al., 2023). Another study of 267 maternal-newborn dyads in Atlanta, GA, reported that maternal PFAS exposures were associated with newborn metabolomic signatures for bioactive lipid metabolism (including leukotrienes), cytochrome p450 pathway, and linoleic acid (Taibl et al., 2023). Collectively, metabolomics studies underscore the importance of lipid metabolism as a potential intermediate mechanism of PFAS exposure.

### 4.3. Strengths and limitations

Our study has notable strengths. First is the diversity of our study population, which included pregnant women from three distinct geographic areas, with high heterogeneity across demographics, socioeconomic status, and PFAS exposures. Second is our statistical approach. We performed combined-cohort analysis utilizing two methods (linear mixed effects models and meta-analysis), and a mixtures analysis using quantile g-computation to assess cumulative associations with PFAS mixtures. The combination of methods applied and the consistency in results across methods strengthens inference in identified associations. Third was our selection of outcomes, which was a targeted assay of bioactive lipids that has not been previously tested for associations with PFAS. This targeted assay directly complements existing studies that have utilized non-targeted metabolomics by deepening knowledge of specific lipid metabolite features to contextualize larger biological pathways and processes observed in past studies. Combined, this resulted in a robust investigation, substantiated by the corroboration of our key findings both within our study and with previous literature.

Our study also has limitations to consider. First is the cross-sectional nature of data collection, as serum PFAS and plasma bioactive lipids were measured during the same visit. Single time point assessment is more susceptible to measurement error and reverse causation than longitudinal studies. Because of their long half-lives, PFAS measures may be relatively stable during pregnancy, however, future studies should consider repeated measures of both PFAS and bioactive lipid levels to reduce measurement error and evaluate more precise windows of vulnerability during pregnancy. Additionally, some confounding variables (e.g. maternal education and parity) were heterogenous across cohorts. There are also unmeasured confounders that we did not model in the present study such as consumer product use and dietary intake which may influence both PFAS and bioactive lipid concentrations. However, exploration of these confounders warrants more comprehensive assessment of the contribution that diet and consumer products have on both PFAS and bioactive lipids, which was beyond the scope of the present study. In terms of statistical approaches, we recognize there are multiple important approaches for analyzing chemical mixtures in health studies. Our study focused on cumulative PFAS mixture associations. However, future studies may consider alternative approaches such as investigation of non-linear effects, high-order interactions across multiple exposure variables, latent clustering of correlated exposure variables, and dimension reduction through development of risk scores or summation based on sources of exposures. Our study evaluated 12 serum PFAS compounds, and there are thousands of PFAS compounds that humans may be exposed to. Therefore, we are likely underestimating the effect of PFAS as a whole class on bioactive lipids. Further, limited sample size and low detection rates of PFAS chemicals in the PROTECT cohort limited our ability to examine differences in associations between Hispanic and White women exposed to PFAS. Additionally, we have previously documented associations between other classes of endocrine disrupting chemicals (e.g., phthalates, phenols, and parabens) and bioactive lipids during pregnancy in the LIFECODES cohort (N = 173) (Aung et al. 2021). It is possible that these chemicals are correlated with prenatal PFAS exposures, contributing to residual confounding and potential interactions, which may influence the effects of PFAS on maternal bioactive lipid profiles. Therefore, future investigations should consider broader chemical mixtures analyses with bioactive lipids and pregnancy outcomes.

### 4.4. Conclusions

In conclusion, we observed associations between PFAS exposure and bioactive lipids during pregnancy in all three enzymatic pathways (cytochrome p450, lipoxygenase, and cyclooxygenase), most of which were positive. Our study complements recent efforts in disentangling endogenous biomarker signatures of PFAS exposures by identifying specific lipid metabolites as biomarkers of exposure and potential mechanistic targets. Future studies should confirm these findings among cohorts of different demographic and socioeconomic makeup, including consideration of meta-analysis and PFAS mixture analysis to strengthen confidence in reported associations. These findings may be utilized to develop hypothesis driven mediation analyses to link PFAS exposures to downstream maternal and child health outcomes. Further, these findings can inform precise risk estimation aimed at reducing the harm of the PFAS exposures on vulnerable populations.

## Abbreviations

CiOB: Chemicals in Our Bodies
CV: coefficient of variation
Et-PFOSA-AcOH: 2-(N-Ethyl-perfluorooctane sulfonamido) acetic acid
IKIDS: Illinois Kids Development Study
Me-PFOSA-AcOH: 2-(N-Methyl-perfluorooctane sulfonamido) acetic acid
NHANES: National Health and Nutrition Examination Study
PFAS: Per- and poly-fluoroalkyl substances
PFBS: Perfluorobutanesulfonic acid
PFDeA: Perfluorodecanoic acid
PFDoA: Perfluorododecanoic acid
PFHpA: Perfluoroheptanoic acid
PFHxS: Perfluorohexane sulfonate
PFNA: Perfluorononanoic acid
PFOA: Perfluorooctanoic acid
PFOS: Perfluorooctanesulfonic acid
PFOSA: Perfluorooctanesulfonamide
PFUdA: Perfluoroundecanoic acid
R^2^: regression coefficients
RSD: relative standard deviation
UIUC: University of Illinois Urbana-Campaign
WQS: weighted quantile sum

## CRediT authorship contribution statement

Funding acquisition (MTA, AA, JDM, JFC, RMF, TJW, SLS, AA); Conceptualization (MTA, HS, AA); Data curation (ED, SN, MW, SME, AC, SP, LZ, JSP, DW, SS, AA, SDG, TJW, RMF, SLS, JDM, AA, MTA, HS); Formal analysis (HS); Investigation (HS, MTA); Methodology (HS, MTA, SME); Literature Review (TM, DP, HS, BAR, MTA); Writing – original draft (HS, MTA); Writing – Reviewing and editing (HS, TM, DP, MW, SME, AC, DJW, RSS, BAR, SP, LZ, DW, JSP, SS, ED, AP, RCF, BM, AA, SDG, SN, GHM, CVV, ZR, JFC, EZ, TJW, RMF, SLS, JDM, AA, MTA).

## Data and code availability

R-script used for the present study are found in supplemental materials.

Select de-identified data from the ECHO Program are available through NICHD’s <u>Data and Specimen Hub (DASH).</u> Information on study data not available on DASH, such as some Indigenous datasets, can be found on the <u>ECHO study DASH webpage.</u>

## Acknowledgements

The authors thank our Environmental Influences on Child Health Outcomes (ECHO) colleagues; the medical, nursing, and program staff; and the children and families participating in the ECHO cohorts. Furthermore, we acknowledge the contribution of the following ECHO program collaborators:

ECHO Components—Coordinating Center: Duke Clinical Research Institute, Durham, North Carolina: Smith PB, Newby LK; Data Analysis Center: Johns Hopkins University Bloomberg School of Public Health, Baltimore, Maryland: Jacobson LP; Research Triangle Institute, Durham, North Carolina: Catellier DJ; Person-Reported Outcomes Core: Northwestern University, Evanston, Illinois: Gershon R, Cella D.

**Appendix A. Supplementary Figure**

**Appendix B. Supplementary Tables**

