## Supplemental Figure 1 for "Cross-sectional associations between prenatal maternal per- and poly-fluoroalkyl substances and bioactive lipids in three Environmental influences on Child Health Outcomes (ECHO) cohorts"

**Supplemental Figure 1**. Flow diagram of final sample selection across CiOB, IKIDS, and PROTECT cohorts

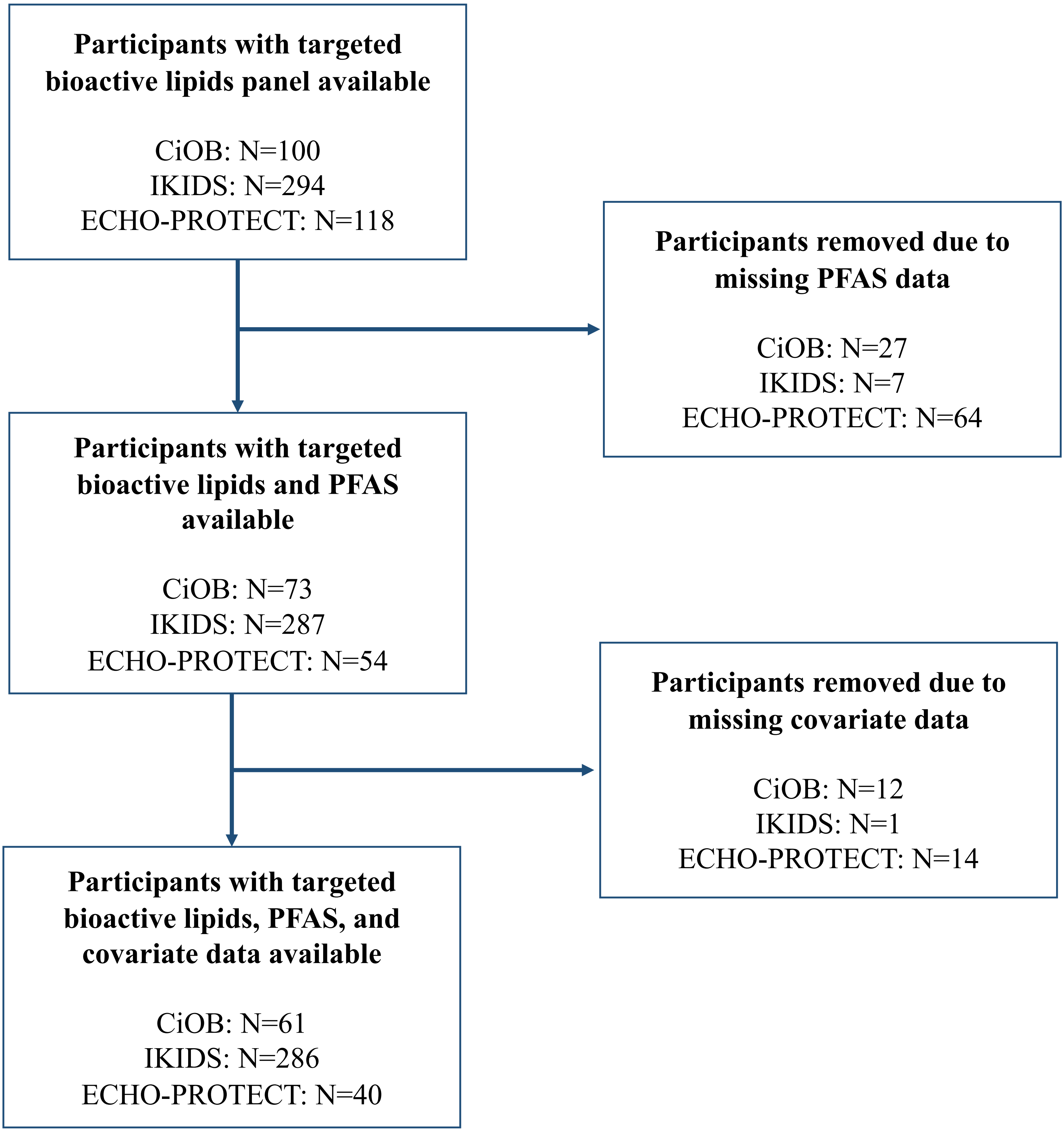
